# Shared Clinical and Immunological Features of Post-Acute COVID-19 and Post-Acute COVID-19 Vaccination Syndromes

**DOI:** 10.64898/2026.09.18.26363380

**Authors:** Johannes Dinser, Moritz Anft, Lea Wiemers, Sarah Skrzypczyk, Eva Kohut, Julia Kurek, Ulrik Stervbo, Felix Seibert, Timm H. Westhoff, Kamil Rosiewicz, Nina Babel

**Affiliations:** Institute for Clinical Immunology, Marien Hospital Herne, University Hospital of the Ruhr-University Bochum, Hölkeskampring 40, 44625 Herne, Germany; Medical Department 1, Marien Hospital Herne, University Hospital of the Ruhr-University Bochum, Hölkeskampring 40, 44625 Herne, Germany; Berlin Institute of Health, Berlin-Brandenburg Center for Regenerative Therapies, and Institute of Medical Immunology, Charité – Universitätsmedizin Berlin, Corporate Member of Freie Universität Berlin, Humboldt-Universität zu Berlin Augustenburger Platz 1, 13353 Berlin, Germany

**Keywords:** Post-COVID, Post-VAC, GPCR-autoantibodies, Immune dysregulation, T-cell autoimmunity

## Abstract

**Background:** Post-acute COVID-19 syndrome (PACS) can cause persistent disability involving fatigue, post-exertional malaise, neurocognitive symptoms, and autonomic manifestations. A clinically overlapping condition has been reported following COVID-19 vaccination and termed post-acute COVID-19 vaccination syndrome (PACVS), but its underlying mechanisms remain poorly understood. We investigated whether immune dysregulation, autoreactivity, antigen persistence, and viral reactivation—mechanisms implicated in PACS—are also present in PACVS.

**Methods:** This cross-sectional study included 28 PACS patients, 29 PACVS patients, and 16 convalescent controls (CCG). Clinical manifestations were assessed using ME/CFS-related symptom questionnaires and the Bell Score. T-cell phenotypes and T cells reactive to SARS-CoV-2 Spike, β1/β2-adrenergic receptor, and M3/M4 muscarinic receptor antigens were analyzed by multicolor flow cytometry. Circulating Spike protein and GPCR autoantibodies were quantified by ELISA, and EBV load by qPCR.

**Results:** PACS and PACVS showed substantial clinical overlap, although several symptoms were more severe in PACS. GPCR autoantibody profiles were comparable between patient groups, while β2-adrenergic receptor autoantibodies were increased compared with CCG. Only three of 57 conventional immunophenotyping parameters differed between PACS and PACVS in unadjusted analyses, whereas both groups showed multiple alterations compared with CCG. Circulating Spike protein was detected in both patient groups, with the highest prevalence in PACVS. β1/β2- and M3/M4-reactive CD4+ T cells were increased, whereas Spike-specific CD4+ T cells were reduced compared with CCG. EBV reactivation was rare.

**Conclusions:** PACS and PACVS exhibit substantial clinical and immunological overlap, including GPCR-directed autoreactivity and altered receptor-reactive T-cell responses. These shared immune-mediated features suggest overlapping pathophysiological mechanisms and warrant investigation of whether therapeutic strategies currently explored for PACS may also be relevant to PACVS.

## 1. Introduction

Post-acute COVID-19 syndrome (PACS), commonly referred to Long COVID, remains an important cause of persistent morbidity following SARS-CoV-2 infection (1). Several definitions have been proposed, differing primarily in the timing and required duration of symptoms. The World Health Organization defines post-COVID-19 condition as symptoms that usually occur within three months after SARS-CoV-2 infection, persist for at least two months, and cannot be explained by an alternative diagnosis (2). Prevalence estimates vary according to study population, viral variant, vaccination status, follow-up period, and case definition, with rates of approximately 10–20% reported in earlier studies (2, 3). PACS can occur following infections of varying severity, and several demographic and clinical factors have been associated with an increased risk, including female sex, smoking, obesity, socioeconomic deprivation, and pre-existing comorbidities (4–6).

A persistent symptom complex clinically resembling PACS has also been reported following SARS-CoV-2 vaccination and has been termed post-acute COVID-19 vaccination syndrome (PACVS) (7). Reported manifestations include fatigue, post-exertional malaise (PEM), cognitive impairment, headache, visual disturbances, palpitations, autonomic dysfunction, and musculoskeletal symptoms (7, 8). In contrast to PACS, PACVS remains insufficiently characterized, epidemiological data are scarce, and standardized diagnostic criteria have not been established (7, 9–11). Consequently, both its prevalence and underlying biological mechanisms remain uncertain.

PACS is a heterogeneous multisystem condition involving fatigue, PEM, cognitive impairment, dyspnea, autonomic dysfunction, palpitations, and muscle and joint pain (4, 8, 12). PACS belongs to the broader spectrum of post-acute infection syndromes, in which myalgic encephalomyelitis/chronic fatigue syndrome (ME/CFS) represents an important clinical manifestation, particularly in patients with persistent fatigue, post-exertional malaise, cognitive impairment, orthostatic syndromes and other multisystem symptoms (4, 8, 12). The pathophysiology of PACS remains incompletely understood and is likely multifactorial (12). Proposed mechanisms include persistent immune dysregulation and inflammation, altered cellular immunity, viral or antigen persistence, reactivation of latent viruses, endothelial or microvascular dysfunction, and autoimmunity (3, 12–22). Persistent alterations in cellular immunity have been described, including changes in naïve and memory T-cell populations and altered expression of activation and exhaustion-associated markers such as PD-1 and TIM-3 (4, 13–16). Increased concentrations of inflammatory mediators, including IL-1β, IL-6, and TNF-α, have additionally been reported (17). These observations suggest that persistent or dysregulated immune responses may contribute to PACS.

Persistence of viral components represents another proposed mechanism. SARS-CoV-2 antigens, including Spike protein, have been detected in subsets of patients months after acute infection, raising the possibility of persistent viral or antigen reservoirs (3, 12, 18–20). Reactivation of latent viruses, particularly Epstein–Barr virus (EBV), has also been associated with PACS and may contribute to immune dysregulation in a subset of patients (4, 13, 15). However, the pathophysiological relevance of viral persistence and reactivation remains under investigation.

Autoimmunity represents another potential mechanism. Different autoantibody specificities have been reported in PACS, including antinuclear antibodies and autoantibodies targeting G-protein-coupled receptors (GPCRs) (13, 20–22). Of particular interest are autoantibodies against β-adrenergic, muscarinic, angiotensin, and endothelin receptors, which are involved in autonomic, vascular, and cardiovascular regulation. Altered immune reactivity against these targets may therefore be relevant to symptoms such as fatigue, orthostatic intolerance, palpitations, and other manifestations of autonomic dysfunction (13, 20–22). Nevertheless, the functional and pathogenic relevance of these autoantibodies remains to be established.

Compared with PACS, considerably less is known about the immunological alterations associated with persistent symptoms following SARS-CoV-2 vaccination. Preliminary studies have reported circulating Spike protein, GPCR-directed autoantibodies, and alterations in inflammatory mediators in individuals with persistent post-vaccination symptoms (9, 23, 24). Autoimmune mechanisms, including molecular mimicry and dysregulated immune responses following antigen exposure, have been proposed, although direct evidence remains limited (25, 26). Reactivation of latent herpesviruses has also been described following vaccination, but its relevance to persistent post-vaccination symptoms remains uncertain (9).

The considerable clinical overlap between PACS and PACVS raises the question of whether these conditions, despite different initiating events, share downstream immunological alterations. Direct comparative studies remain scarce, and it is unclear whether PACS and PACVS share changes in conventional T-cell phenotypes, autoreactivity against receptors involved in autonomic regulation, receptor-reactive T-cell responses, persistence of circulating Spike protein, or evidence of latent viral reactivation.

The present cross-sectional study therefore aimed to directly compare the clinical and immunological characteristics of patients with PACS and PACVS. We assessed symptom profiles and functional impairment, performed comprehensive peripheral T-cell immunophenotyping, quantified circulating Spike protein and GPCR-directed autoantibodies, characterized CD4+ and CD8+ T-cell responses to SARS-CoV-2 Spike and β1/β2-adrenergic and M3/M4 muscarinic receptor antigens, and assessed EBV load. By comparing PACS and PACVS with each other and with convalescent controls, we aimed to identify shared clinical and immunological characteristics as well as features distinguishing the two conditions.

## 2. Materials and Methods

### 2.1 Study population and study design

This cross-sectional study included 57 adult randomly selected patients previously diagnosed with either post-acute COVID-19 syndrome (PACS; n = 28) or post-acute COVID-19 vaccination syndrome (PACVS; n = 29). Patients were recruited at the Center for Translational Medicine, Marien Hospital Herne, Germany. In addition, a convalescent control group (CCG) comprising 16 individuals with a history of SARS-CoV-2 infection and SARS-CoV-2 vaccination was included.

Eligibility criteria for the patient cohorts were age ≥18 years, a diagnosis of PACS or PACVS, and written informed consent. Blood samples obtained as part of routine diagnostic procedures were used for the analyses. Because individual assays were implemented at different time points during the study, not all measurements were available for every participant. Consequently, sample sizes differed between individual analyses and are reported where applicable.

The study was approved by the Ethics Committee of the Medical Association of Westphalia-Lippe and the University of Münster (reference number 2024-639-f-S). Written informed consent was obtained from all study participants prior to inclusion in the study.

### 2.2 Blood collection and sample processing

Peripheral venous blood was collected into S-Monovette K3 EDTA and S-Monovette Serum tubes (Sarstedt, Germany) and processed on the following day.

For isolation of peripheral blood mononuclear cells (PBMCs), 1.5 mL of EDTA-anticoagulated blood was diluted 1:1 with phosphate-buffered saline (PBS; PAN-Biotech GmbH, Germany) and layered onto 15 mL Pancoll density-gradient medium (PAN-Biotech GmbH, Germany). Samples were centrifuged at 800 × g for 20 min at room temperature. PBMCs were collected and washed twice with PBS. Cells were subsequently resuspended in fetal calf serum (FCS; PAN-Biotech GmbH, Germany) containing dimethyl sulfoxide (DMSO; Sigma-Aldrich, USA) and stored at −80 °C until further analysis.

Serum samples were centrifuged at 2,500 × g for 10 min at 4 °C. Serum was subsequently aliquoted and stored at −80 °C until analysis.

### 2.3 Immunophenotyping

For conventional immunophenotyping, optimized concentrations of fluorochrome-conjugated antibodies were added directly to EDTA-anticoagulated whole blood and incubated for 15 min at room temperature in the dark. Erythrocytes were subsequently lysed using VersaLyse (Beckman Coulter, USA) for 30 min at room temperature in the dark. Samples were washed with PBS and analyzed using a CytoFLEX flow cytometer (Beckman Coulter, USA). The gating strategies are shown in Supplementary *Figures S4–S7*.

### 2.4 Antigen-specific T-cell stimulation

Cryopreserved PBMCs were thawed and washed with RPMI medium (Gibco, USA) supplemented with 10% FCS and 2% penicillin-streptomycin-glutamine (Sigma-Aldrich, USA). Cells were incubated at 37 °C before antigen-specific stimulation.

PBMCs were stimulated separately with 4 µL SARS-CoV-2 Spike protein antigen (Miltenyi Biotec, Germany), 4 µL cell lysates containing muscarinic acetylcholine receptors M3 and M4 (Biotrend, Germany), or 4 µL cell lysates containing β1- and β2-adrenergic receptors (Biotrend, Germany). Optimal concentrations for stimulation with the receptor cell lysates were determined beforehand by titration. Separate PBMC aliquots were stimulated with 0.2 µL staphylococcal enterotoxin B (SEB; Sigma-Aldrich, USA) as a positive control.

After 2 h of stimulation at 37 °C, 0.4 µL Brefeldin A (Sigma-Aldrich, USA) was added. Cells were subsequently incubated for a total stimulation period of 18 h before antibody staining and multicolor flow-cytometric analysis. The gating strategies used for antigen-specific T-cell analyses are shown in Supplementary *Figures S2* and *S3*.

### 2.5 Flow cytometry of antigen-specific T cells

Following antigen stimulation, PBMCs underwent extracellular surface staining with the respective antibodies as described in 2.6. for 10 min at room temperature in the dark. Cells were washed twice with PBS containing bovine serum albumin (PBS/BSA) and subsequently fixed and permeabilized using the Intracellular Fixation & Permeabilization Buffer Set (Thermo Fisher Scientific, USA) according to the manufacturer’s instructions.

Optimized concentrations of antibodies targeting intracellular antigens were added, followed by incubation for 30 min at room temperature in the dark. Samples were analyzed using a CytoFLEX flow cytometer (Beckman Coulter, USA). Daily instrument quality control was performed using CytoFLEX Daily QC Fluorospheres (Beckman Coulter, USA) according to the manufacturer’s recommendations. Flow-cytometry data were analyzed and gated using FlowJo version 10.10.0.

### 2.6 Antibodies

For basic immune-status immunophenotyping, all antibodies were obtained from Beckman Coulter, USA: CD14-FITC (clone RMO52), CD56-PE (clone N901), CD4-ECD (clone SFCI12T4D11), CD19-PC5.5 (clone J3-119), CD8-APC (clone B9.11), CD3-APC-A750 (clone UCHT1), and CD45-PB (clone J33).

For α/β T-memory and regulatory T-cell phenotyping, antibodies included CD127-FITC (clone R34.34), TCRαβ-PE (clone IP26A), CD45RA-ECD (clone 2H4LDH11LDB9), TCRγδ-PC5.5 (clone IMMU510), CCR7-PC7 (clone G043H7), CD4-APC (clone 13B8.2), CD8-APC-A700 (clone B9.11), CD3-APC-A750 (clone UCHT1), CD25-SN428 (clone B1.49.9), and CD45-KrO (clone J33) (all Beckman Coulter, USA).

For T-cell activation phenotyping, antibodies included CD11a-FITC (clone 25.3), HLA-DR-PE (clone Immu-357), CD4-ECD (clone SFCI12T4D11), CD28-PC5.5 (clone CD28.2), CD8-APC (clone B9.11), CD3-APC-A750 (clone UCHT1), and CD57-PB (clone NC1) (all Beckman Coulter, USA).

For antigen-specific T-cell phenotyping, antibodies were obtained from BioLegend, USA, unless otherwise stated. Extracellular staining included CCR7-PerCP-Cy5.5 (clone G043H7), CD4-AF700 (clone OKT4), CXCR5-PE-Dazzle594 (clone MOPC-21), viability dye eFluor780 (Thermo Fisher Scientific, USA), CD8-V500 (clone RPA-T8; BD Biosciences, USA), PD-1-PE (clone A17188A), TIM-3-VioBright FITC (clone REA635; Miltenyi Biotec, Germany), CTLA-4-BV421 (clone BNI3), LAG-3-BV650 (clone 11C3C65), and CD45RA-BV605 (clone HI100). Intracellular staining included CD137-PE-Cy7 (clone 4B4-1), CD154-AF647 (clone 24-31), and CD3-BV785 (clone OKT3).

### 2.7 EBV detection by quantitative real-time PCR

DNA was extracted from peripheral venous blood samples using the AltoStar AM16 automated pipetting system and the AltoStar Purification Kit 1.5 (altona Diagnostics GmbH, Germany). Epstein–Barr virus (EBV) DNA was quantified using the AltoStar EBV PCR Kit 1.5 (altona Diagnostics GmbH, Germany) according to the manufacturer’s instructions.

Quantitative real-time PCR was performed using a Bio-Rad C1000 Thermal Cycler (Bio-Rad, USA). The detection threshold was defined as the lowest viral load within the linear range of the assay (250 copies/mL).

### 2.8 Spike protein ELISA

Serum samples were collected and stored at −80 °C as described above. Circulating SARS-CoV-2 Spike protein was quantified using the Human SARS-CoV-2 RBD ELISA Kit (Thermo Fisher Scientific, USA) according to the manufacturer’s instructions. Optical density was measured using an Asys UVM 340 microplate reader.

### 2.9 GPCR autoantibody ELISA

Serum samples were stored at −80 °C until analysis. Autoantibodies targeting angiotensin II receptor type 1 (AT-II), β1-adrenergic receptor (β1), β2-adrenergic receptor (β2), endothelin A receptor (ET-A), muscarinic acetylcholine receptor M3 (M3), and muscarinic acetylcholine receptor M4 (M4) were quantified using commercially available ELISA systems (CellTrend, Germany) according to the manufacturer’s instructions.

The following cut-off concentrations as provided by manufacturer were used to define autoantibody positivity: AT-II ≥17.0 U/mL, β1 ≥15.0 U/mL, β2 ≥14.0 U/mL, ET-A ≥17.0 U/mL, M3 ≥10.0 U/mL, and M4 ≥10.7 U/mL.

### 2.10 Clinical questionnaires

Clinical manifestations and functional impairment were assessed using three patient-reported questionnaires that had been completed as part of the patients’ previous diagnostic assessment: the Canadian Consensus Criteria (CCC) questionnaire for myalgic encephalomyelitis/chronic fatigue syndrome (ME/CFS), the Bell Disability Scale, and a Subjective Symptom Severity Questionnaire (SSSQ).

The CCC questionnaire assesses ME/CFS-associated manifestations across seven symptom domains, including fatigue, sleep disturbances, pain, neurological/cognitive manifestations, autonomic manifestations, neuroendocrine manifestations, and immune manifestations.

The Bell Disability Scale assesses disease-associated functional impairment on a scale ranging from 0 to 100 in increments of 10, with lower scores indicating greater impairment and 100 indicating absence of symptoms at rest and normal functional capacity.

The SSSQ assesses the severity of symptoms across domains including fatigue, pain, cognitive impairment, circulatory symptoms, sleep disturbances, photosensitivity, and noise sensitivity. Individual symptoms were rated on a numerical scale ranging from 1 (no symptoms) to 10 (most severe symptoms).

### 2.11 Statistical analysis

Statistical analyses were performed using GraphPad Prism version 10.4.1. No data transformation was applied. Continuous variables with non-normally distributed values were compared between two independent groups using the Mann–Whitney U test. Differences in categorical proportions were assessed using Fisher’s exact test where appropriate, and differences in sex distribution were assessed using the chi-squared test. Age differences between groups were analyzed using Welch’s t-test.

Violin plots display the median and first and third quartiles. Box plots display the median. All statistical tests were two-sided, and a p-value <0.05 was considered statistically significant. Because the study was exploratory, p-values were not adjusted for multiple comparisons.

### 2.12 Statistical analysis of disease duration, age and gender effects

To account for differences in disease duration, age and gender between PACS and PACVS, all analyses were repeated using regression models including disease duration (continuous), group (PACS vs. PACVS), and a group-by-disease duration interaction term, adjusted for age and sex. Continuous outcomes were analyzed using linear regression models, whereas binary symptom outcomes were analyzed using logistic regression models. Outcomes were log-transformed where appropriate to improve distributional properties. Model coefficients were extracted for main effects and interaction terms. To account for multiple testing, p-values were adjusted using the Benjamini-Hochberg false discovery rate (FDR) method. Model-based predicted values and 95% confidence intervals were derived using the *ggeffects* package.

## 3. Results

### 3.1 Demographic and clinical characteristics

A total of 57 patients with post-acute COVID-19 syndrome (PACS; n = 28) or post-acute vaccination syndrome (PACVS; n = 29), together with 16 convalescent controls (CCG), were included in the study. All patients in the PACS and PACVS cohorts fulfilled the ME/CFS criteria used in the study.

In the PACS cohort, 25 of 28 patients were female (89.3%), with a median age of 50 years. In the PACVS cohort, 16 of 29 patients were female (55.2%), with a median age of 37 years. Among convalescent controls, 10 of 16 individuals were female (62.5%), with a median age of 36 years (Supplementary Information *Table S1*).

The study groups differed significantly with respect to sex and age. The proportion of female participants was significantly higher in PACS than in PACVS (p = 0.0042) and CCG (p = 0.0341), whereas no significant difference was observed between PACVS and CCG (p = 0.6338; Chi-squared test). PACS patients were also significantly older than both PACVS patients (p = 0.0146) and convalescent controls (p = 0.0124), while age did not differ significantly between PACVS and CCG (p = 0.5263; Welch’s t-test).

The interval between antigen exposure and immunological assessment was significantly longer in PACVS than in PACS across the investigated analyses (Supplementary Information *Table S2*). One PACS patient was excluded from this analysis because the exact date of antigen exposure was unknown.

### 3.2 PACS and PACVS show substantial clinical overlap, with greater severity of selected symptoms in PACS

Clinical manifestations were assessed using three patient-reported outcome measures addressing symptom prevalence, symptom severity, and disease-associated functional impairment.

The Canadian Consensus Criteria questionnaire, assessing 38 ME/CFS-associated symptoms, was completed by 27 PACS and 26 PACVS patients. Post-exertional malaise and cognitive impairment were among the most frequently reported symptoms in both cohorts.

Dizziness/lightheadedness was significantly more prevalent in PACS than in PACVS (p = 0.01135, Fisher’s exact test), whereas no significant between-group differences were observed for the remaining symptoms assessed by this questionnaire.

Disease-associated functional impairment was evaluated using the Bell Score in 27 PACS and 29 PACVS patients (Figure 1A). Mean Bell Scores were 34.63 ± 15.56 in PACS and 39.31 ± 14.06 in PACVS. The difference between the groups was not statistically significant, demonstrating a comparable overall degree of functional impairment, with both cohorts showing moderate-to-severe limitations in daily activities.

**Figure 1:**
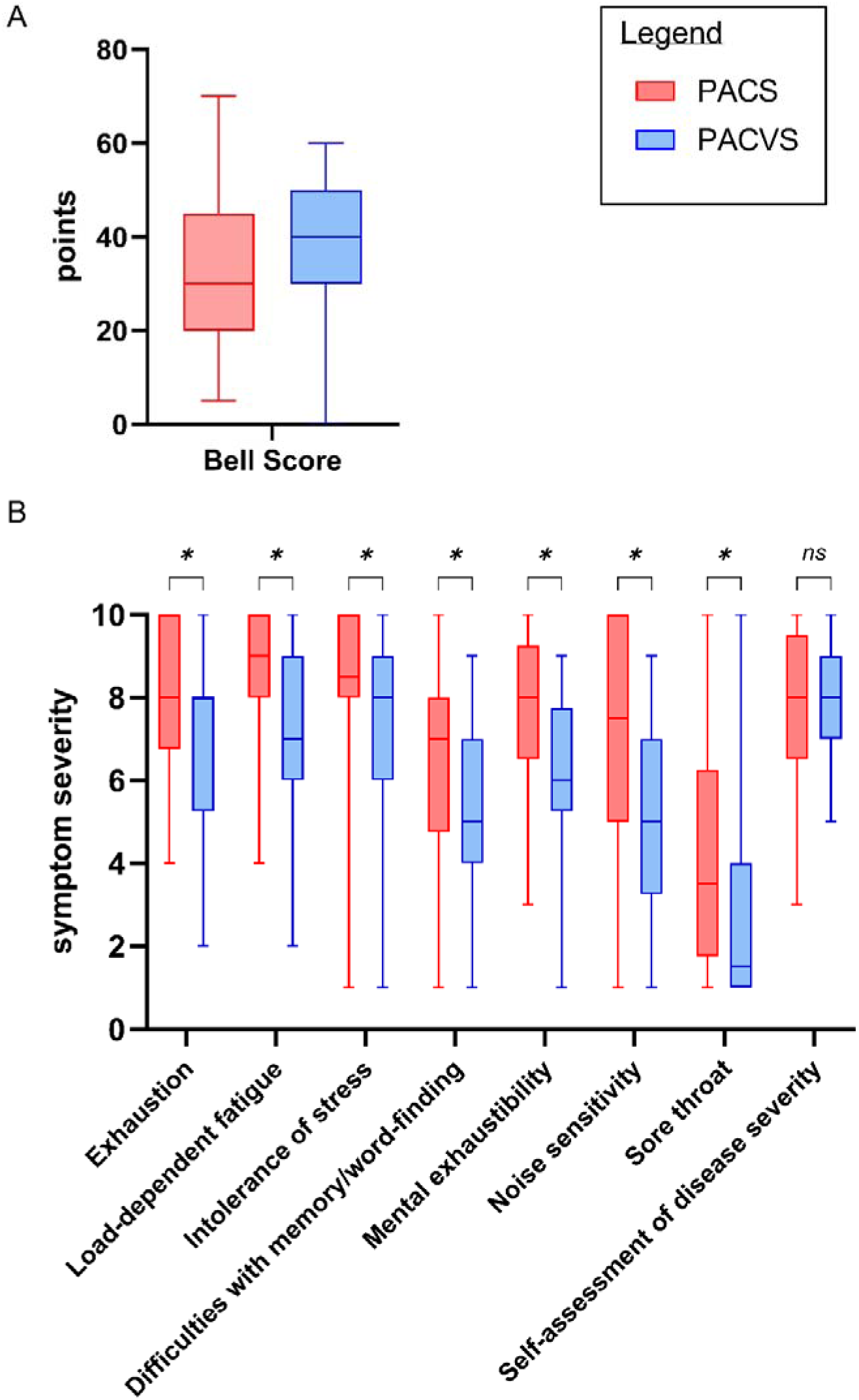
**(A)**: Comparison of subjective Bell Score quantifying impairments in daily life with 0 points = most severe impairments and 100 points = no impairments. **(B)**: Questionnaire used for quantification of symptom severity (SSSQ) with 0 points = no symptoms and 10 points = most severe symptoms. Statistical analysis was performed using Fisher’s exact test with the following p-values: * = p<0.05.

Individual symptom severity was further assessed using the Subjective Symptom Severity Questionnaire (SSSQ) (Figure 1B). Seven symptoms were significantly more severe in PACS than in PACVS: exhaustion, load-dependent fatigue, intolerance of stress, memory or word-finding difficulties, mental exhaustibility, noise sensitivity, and sore throat.

Thus, despite substantial overlap in the clinical phenotype and comparable overall functional impairment, PACS patients experienced greater severity of selected symptoms than PACVS patients.

### 3.3 PACS and PACVS show largely overlapping T-cell immunophenotypes

Comprehensive peripheral blood immunophenotyping comprised 57 parameters. Only three of these parameters differed significantly between PACS and PACVS (Figure 2). PACS patients exhibited a higher proportion of CD45RA− memory CD4+ T cells (p = 0.019591), a lower proportion of naïve CD8+ T cells (CD45RA+CCR7+; p = 0.039191), and a higher proportion of HLA-DR+ CD4+ T cells (p = 0.008646) compared with PACVS patients (Mann–Whitney U-test).

**Figure 2:**
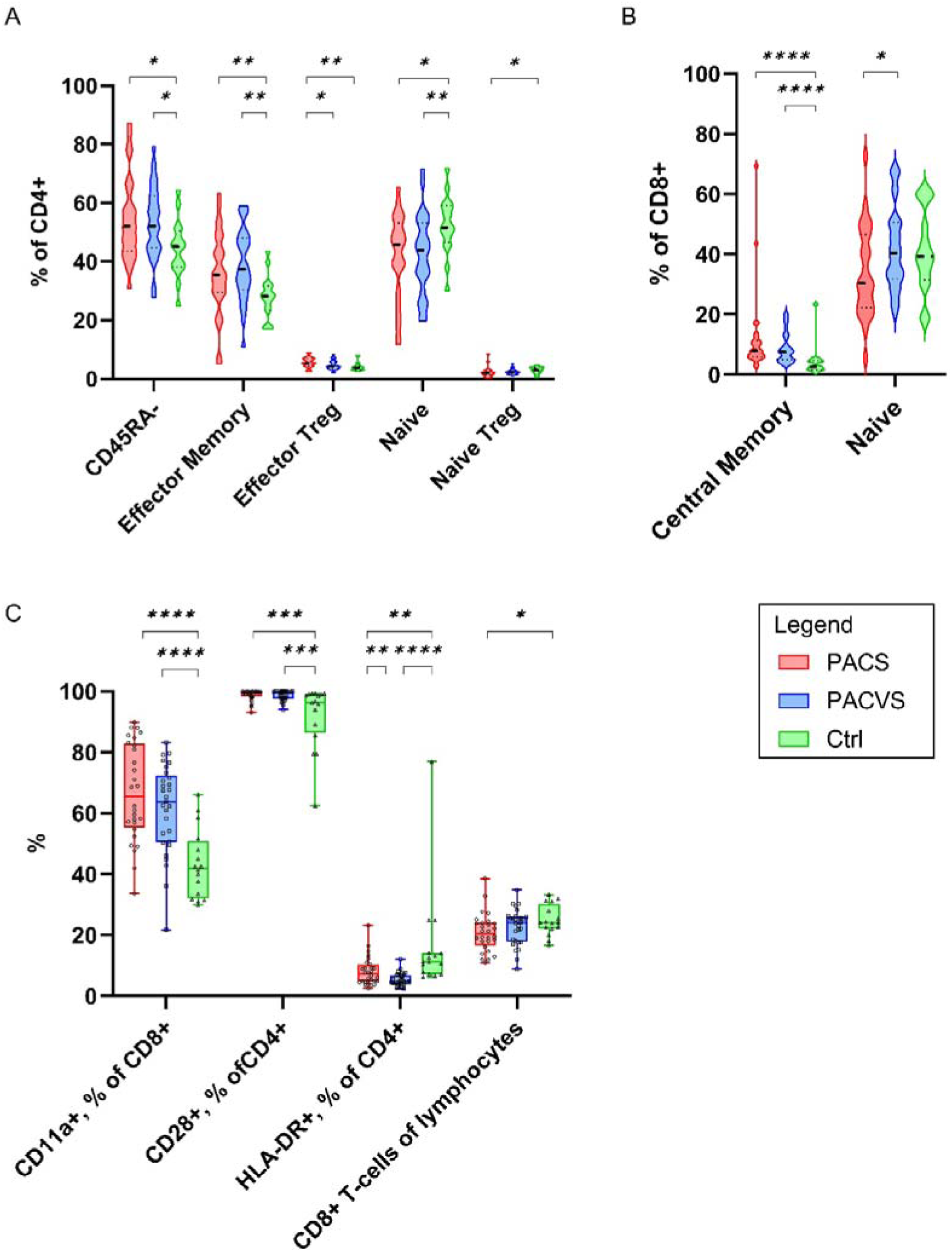
**(A)/(B)**: Immunophenotyping analysis, significant differences of the subgroup α/β TmemTreg. Statistical analysis performed was Mann-Whitney U-test with the following p-values: * = p<0.05, ** = p<0.01, *** = p<0.001, **** = p<0.0001. **(C)**: Immunophenotyping analysis, significant differences of the subgroup TC act and CD8^+^ T-cells of lymphocytes. Statistical analysis performed was Mann-Whitney U-test with the following p-values: * = p<0.05, ** = p<0.01, *** = p<0.001, **** = p<0.0001.

More pronounced differences were observed when PACS and PACVS were compared with CCG. Within the CD4+ compartment, both patient groups showed higher proportions of CD45RA− cells (PACS vs. CCG: p = 0.015061; PACVS vs. CCG: p = 0.010447) and effector-memory cells (PACS vs. CCG: p = 0.008301; PACVS vs. CCG: p = 0.001208). Conversely, naïve CD4+ T cells (CD45RA+CCR7+) were more frequent in CCG than in PACS (p = 0.015061) or PACVS (p = 0.006653). Naïve regulatory T cells were additionally increased in CCG compared with PACS (p = 0.024809) (Figure 2A).

Within the CD8+ compartment, central-memory T cells were significantly more frequent in both PACS and PACVS than in CCG (PACS vs. CCG: p = 0.000002; PACVS vs. CCG: p = 0.000031) (Figure 2B). Expression of CD11a was increased in PACS and PACVS compared with CCG (PACS vs. CCG: p = 0.000009; PACVS vs. CCG: p = 0.000052), as was CD28 expression (PACS vs. CCG: p = 0.000353; PACVS vs. CCG: p = 0.000974). In contrast, HLA-DR expression was lower in PACS (p = 0.003915) and PACVS (p < 0.000001) compared with CCG. PACS additionally exhibited a lower proportion of CD8+ T cells among total lymphocytes than CCG (p = 0.018901) (Figure 2C).

Overall, conventional T-cell phenotypes showed substantial overlap between PACS and PACVS, whereas both patient cohorts exhibited multiple alterations compared with convalescent controls.

### 3.4 GPCR autoantibody profiles are comparable between PACS and PACVS

Autoantibodies targeting six G-protein-coupled receptors were quantified: angiotensin II receptor type 1 (AT-II), β1- and β2-adrenergic receptors, endothelin A receptor (ET-A), and muscarinic acetylcholine receptors M3 and M4.

Using predefined cut-off concentrations, PACS and PACVS showed broadly comparable patterns of GPCR autoantibody positivity (Figure 3A). Autoantibodies against β-adrenergic receptors were among the most frequently detected autoantibodies in both patient cohorts. Convalescent controls generally showed a lower prevalence of detectable GPCR autoantibodies, with a significant difference observed for β2-adrenergic receptor autoantibody positivity.

**Figure 3:**
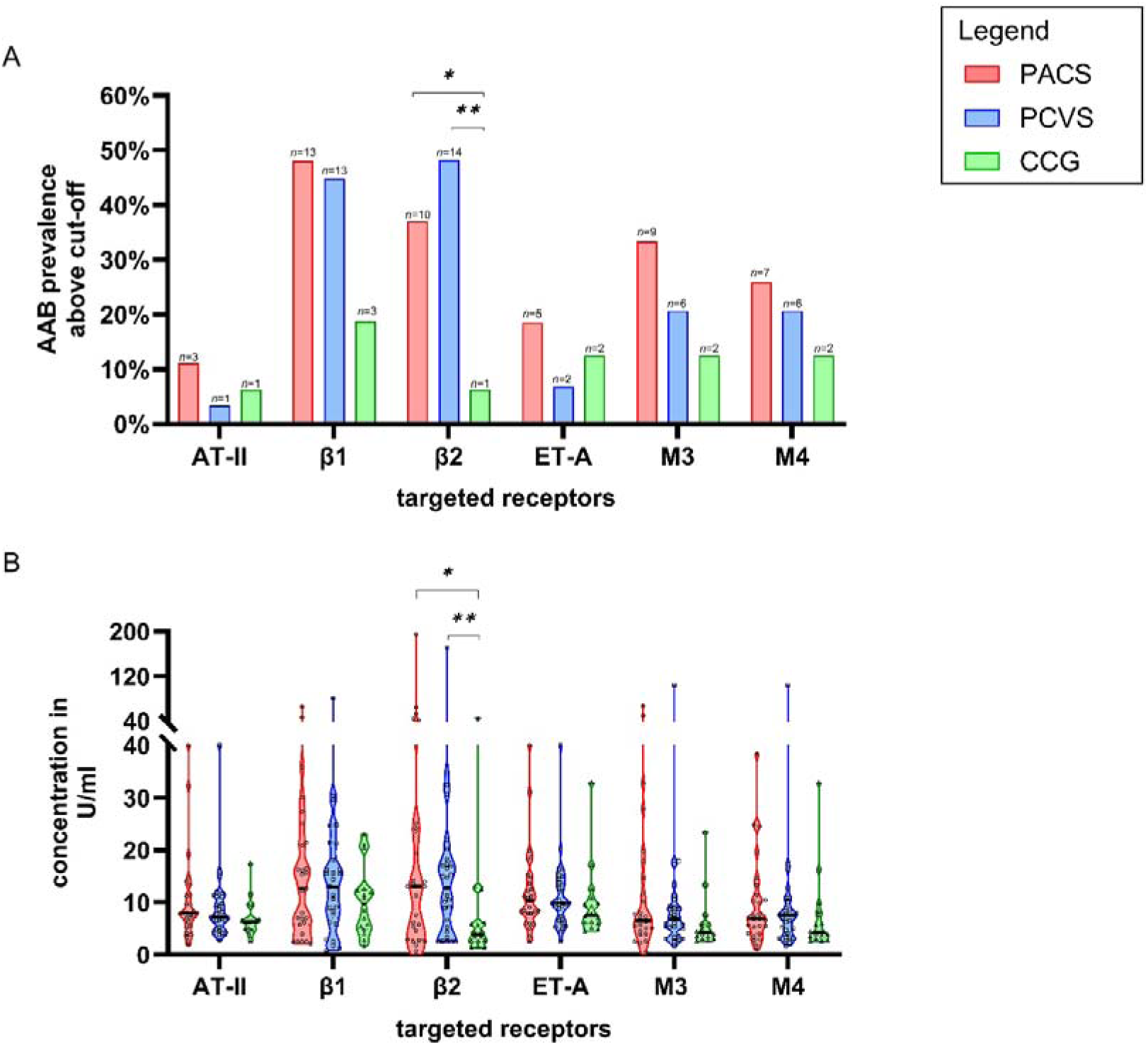
**(A)**: Prevalence of autoantibodies above cut-off values. Autoantibodies above the cut-off are considered as a positive autoantibody status. Evaluation of differences between the groups was done using Fisher’s exact test with * = p<0.05 and ** = p<0.01. **(B)**: Concentration of autoantibodies in PACS and PACVS patients compared to CCG. Higher levels of autoantibodies targeting β2-receptors occur in both patient groups. Intergroup differences were assessed using the Mann-Whitney U-Test with * = p<0.05 and ** = p<0.01.

No significant differences in GPCR autoantibody concentrations were observed between PACS and PACVS (AT-II: p = 0.6991; β1: p = 0.9968; β2: p = 0.9061; ET-A: p = 0.9255; M3: p = 0.8740; M4: p = 0.7112) (Figure 3B).

In contrast, β2-adrenergic receptor autoantibody concentrations were significantly increased in both PACS and PACVS compared with CCG (PACS vs. CCG: p = 0.0246; PACVS vs. CCG: p = 0.007) (Figure 3B).

These findings indicate highly overlapping GPCR autoantibody profiles in PACS and PACVS, while increased β2-adrenergic receptor autoantibody concentrations distinguish both symptomatic patient cohorts from convalescent controls.

### 3.5 Circulating Spike protein is detectable in PACS and PACVS and is most prevalent in PACVS

Soluble circulating Spike protein was quantified by ELISA. Serum samples were available from 25 of 28 PACS patients, 28 of 29 PACVS patients, and all convalescent controls.

Spike protein was detected in 6 of 25 PACS patients (24.0%), 12 of 28 PACVS patients (42.86%), and 2 of 16 convalescent controls (12.5%) (Figure 4A). Although Spike positivity was numerically more frequent in PACVS than in PACS, this difference did not reach statistical significance.

**Figure 4:**
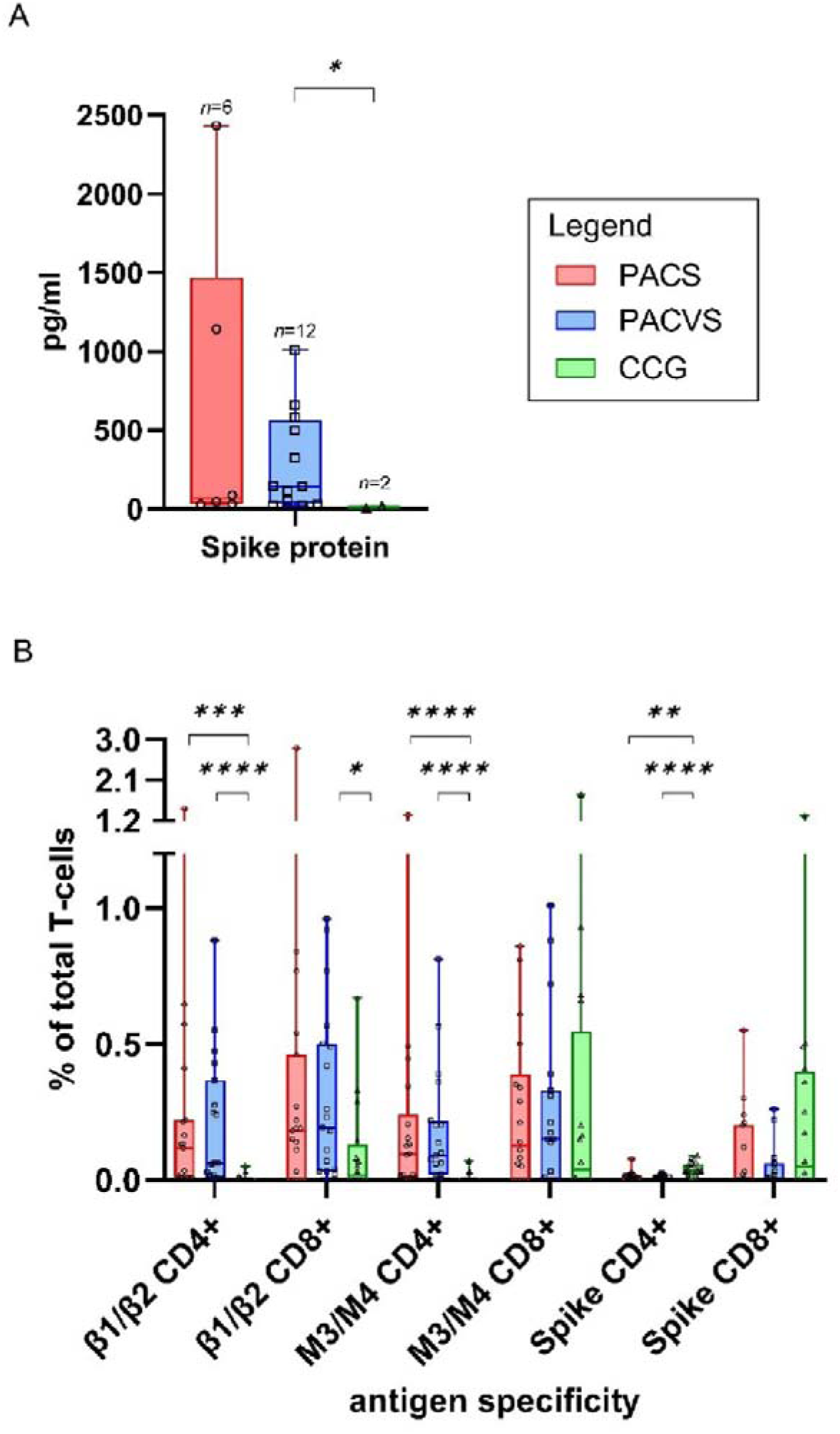
**(A)**: Comparison of the positive Spike protein titers in PACS, PACVS and convalescent control patients. Statistical analysis used: Mann-Whitney U-test, * = p<0.05, n=number. **(B)**: Comparison of β1/β2-, M3/M4-AABs and SARS-CoV-2 Spike protein specific T-cells in PACS, PACVS and convalescent control patients. CD4 T cells expressing CD154 and CD137 and CD8 T cells expressing CD137 were defined as antigen-specific T cells. Statistical analysis was performed using Mann-Whitney U-test with * = p<0.05, ** = p<0.01, *** = p<0.001, **** = p<0.0001.

Quantitative analysis showed higher circulating Spike concentrations in PACS and PACVS than in CCG, the later showed almost neglectable Spike concentration (Figure 4A).

Thus, circulating Spike protein was detectable in subsets of both patient cohorts, with the highest numerical prevalence observed in PACVS.

### 3.6 PACS and PACVS exhibit increased CD4+ T-cell responses to β-adrenergic and muscarinic receptor antigens

Antigen-specific T-cell responses against β1/β2-adrenergic receptor cell lysates, M3/M4 muscarinic receptor cell lysates, and SARS-CoV-2 Spike overlapping peptide pools were assessed. Antigen-specific CD4+ T cells were identified by co-expression of CD137 and CD154, whereas antigen-specific CD8+ T cells were defined by CD137 expression.

β1/β2-, M3/M4-, and Spike-specific T cells were detectable in both PACS and PACVS. Direct comparison revealed no significant differences in the overall frequencies of these antigen-specific T-cell populations between the two patient cohorts.

In contrast, β1/β2-specific CD4+ T-cell frequencies were significantly increased in PACS (p = 0.000175) and PACVS (p = 0.000016) compared with CCG. M3/M4-specific CD4+ T-cell frequencies were likewise increased in PACS (p = 0.000065) and PACVS (p = 0.000017) (Figure 4B).

Spike-specific CD4+ T cells showed the opposite pattern, with significantly lower frequencies in PACS (p = 0.001466) and PACVS (p = 0.000073) than in CCG. β1/β2-specific CD8+ T cells were additionally increased in PACVS compared with CCG (p = 0.014952) (Figure 4B).

These findings demonstrate a shared pattern of antigen-specific cellular immunity in PACS and PACVS, characterized by increased CD4+ T-cell reactivity against β-adrenergic and muscarinic receptor-derived antigens relative to convalescent controls.

### 3.7 Phenotypic alterations predominantly affect receptor-specific CD4+ T cells

Given the increased frequencies of β1/β2- and M3/M4-specific CD4+ T cells in both PACS and PACVS compared with CCG, we next investigated whether these antigen-specific T-cell populations also exhibited distinct phenotypic characteristics. To this end, antigen-specific T cells were further characterized with respect to CTLA-4, LAG-3, PD-1, TIM-3 and CXCR5 expression, together with the distribution of naïve, central-memory (CM), effector-memory (EM), and TEMRA subsets.

The most pronounced differences between symptomatic patients and CCG occurred among β1/β2- and M3/M4-specific CD4+ T cells (Table 1). Both PACS and PACVS showed multiple significant differences in CTLA-4, CXCR5, LAG-3 and, for selected comparisons, PD-1 expression, together with alterations across naïve, CM, EM, and TEMRA compartments.

**Table 1:** Display of differences in cell type and surface markers between PACS, PACVS and convalescent control patients (CCG). The numbers represent *p*-values of the Mann-Whitney U-test, bold values are considered significant. Numbers written in the color red indicate a higher percentage of PACS compared to other, numbers written in the color blue indicate a higher percentage of PACVS compared to other, numbers written in green indicate a higher percentage of CCG compared to other.

| PACS/CCG | CTLA-4<sup>+</sup> | CXCR-5<sup>+</sup> | Lag-3<sup>+</sup> | PD-1<sup>+</sup> | TIM-3<sup>+</sup> | Naive TC | CM | EM | TEMRA |
| --- | --- | --- | --- | --- | --- | --- | --- | --- | --- |
| β1/β2 CD4<sup>+</sup> | **0.0185** | **<0.0001** | **0.0177** | 0.0662 | 0.6318 | **0.0005** | **0.0001** | **0.0136** | **0.0334** |
| β1/β2 CD8<sup>+</sup> | 0.8811 | 0.4337 | 0.3599 | 0.9129 | 0.8235 | 0.2174 | 0.2226 | 0.0551 | 0.2441 |
| M3/M4 CD4<sup>+</sup> | **0.002** | **0.0015** | **0.0177** | **0.0411** | 0.8793 | **0.0004** | **<0.0001** | **0.0171** | **0.0144** |
| M3/M4 CD8<sup>+</sup> | 0.8355 | 0.7124 | 0.1524 | 0.7137 | 0.7121 | 0.9649 | 0.7568 | **0.0355** | 0.3694 |
| Spike CD4<sup>+</sup> | 0.9094 | 0.8086 | 0.0897 | 0.3264 | 0.0638 | 0.6356 | 0.7847 | **0.0263** | 0.2995 |
| Spike CD8<sup>+</sup> | 0.3787 | 0.3917 | 0.4681 | 0.2035 | 0.1248 | **0.0411** | 0.2312 | 0.6383 | 0.4004 |

| PACVS/CCG | CTLA-4<sup>+</sup> | CXCR-5<sup>+</sup> | Lag-3<sup>+</sup> | PD-1<sup>+</sup> | TIM-3<sup>+</sup> | Naive TC | CM | EM | TEMRA |
| --- | --- | --- | --- | --- | --- | --- | --- | --- | --- |
| β1/β2 CD4<sup>+</sup> | **0.0009** | **<0.0001** | **0.0036** | **0.0308** | >0.9999 | **<0.0001** | **<0.0001** | **0.0025** | **0.0069** |
| β1/β2 CD8<sup>+</sup> | 0.6991 | **0.0276** | 0.0911 | 0.9034 | 0.3842 | **0.0065** | 0.0587 | **0.0157** | 0.0817 |
| M3/M4 CD4<sup>+</sup> | **<0.0001** | **0.0003** | **0.0108** | **0.0024** | 0.9688 | **0.0002** | **<0.0001** | **0.0065** | 0.0889 |
| M3/M4 CD8<sup>+</sup> | 0.9825 | 0.9598 | 0.1919 | 0.8779 | 0.8779 | 0.7821 | 0.8459 | 0.0741 | 0.3376 |
| Spike CD4<sup>+</sup> | 0.5944 | 0.6689 | 0.4483 | **0.007** | 0.1077 | 0.6244 | 0.8662 | **0.01** | 0.1724 |
| Spike CD8<sup>+</sup> | 0.1929 | 0.6226 | 0.6495 | 0.1494 | 0.1494 | 0.9933 | 0.4172 | 0.3955 | 0.3309 |

| PACS/PACVS | CTLA-4<sup>+</sup> | CXCR-5<sup>+</sup> | Lag-3<sup>+</sup> | PD-1<sup>+</sup> | TIM-3<sup>+</sup> | Naive TC | CM | EM | TEMRA |
| --- | --- | --- | --- | --- | --- | --- | --- | --- | --- |
| β1/β2 CD4<sup>+</sup> | 0.7771 | 0.3066 | 0.4079 | 0.7166 | 0.6855 | 0.8795 | 0.9597 | 0.2736 | 0.3302 |
| β1/β2 CD8<sup>+</sup> | 0.0846 | 0.0646 | 0.5153 | 0.8465 | **0.0440** | **0.0339** | 0.0854 | 0.6639 | 0.7656 |
| M3/M4 CD4<sup>+</sup> | 0.6910 | 0.7569 | 0.6160 | 0.4321 | 0.7712 | 0.9102 | 0.8276 | **0.5019** | 0.1808 |
| M3/M4 CD8<sup>+</sup> | 0.4116 | 0.6852 | 0.9369 | 0.5848 | 0.3788 | 0.5938 | 0.8367 | 0.5371 | 0.9877 |
| Spike CD4<sup>+</sup> | 0.7379 | 0.9947 | 0.5956 | 0.2245 | 0.6630 | 0.9477 | 0.5395 | 0.3395 | 0.1197 |
| Spike CD8<sup>+</sup> | 0.7088 | 0.6387 | 0.9309 | 0.6321 | 0.9525 | 0.1395 | 0.6230 | 0.8853 | 0.9589 |

In contrast, direct PACS–PACVS comparisons revealed very few significant differences. Among β1/β2-specific CD8+ T cells, PACS and PACVS differed in TIM-3 expression (p = 0.0440) and the proportion of naïve cells (p = 0.0339). No significant PACS–PACVS differences were detected in the assessed phenotypic parameters of β1/β2-specific CD4+, M3/M4-specific CD4+ or CD8+, or Spike-specific CD4+ or CD8+ T cells.

### 3.8 Disease duration, sex and age have limited influence on the observed immune phenotype

Because disease duration differed significantly between PACS and PACVS, all analyses were repeated using regression models adjusted for disease duration, age, and sex, and extended by group-by-disease-duration interaction terms. Across standard laboratory parameters, autoantibodies, soluble Spike protein, ex vivo immunophenotyping, T-cell stimulation assays, and cognitive symptoms scores, no robust effects of disease duration or group-by-duration interaction were detected after correction for multiple testing.

For binary symptom data (CCC), logistic regression analyses revealed no significant associations between disease duration, group, or their interaction and the presence of any reported symptoms after False Discovery Rate (FDR) correction. Similarly, continuous symptom severity scores (SSSQ) analyzed using linear regression models showed no significant effects after multiple testing correction. Although several nominal associations were observed at the unadjusted level, none remained significant after FDR correction.

In baseline immunophenotyping, one significant interaction remained after FDR correction for central memory CD45RA-CCR7+ CD8+ T cells. Model-based predictions showed a pronounced decline of this subset with increasing disease duration in PACS, whereas the trajectory in PACVS was comparatively stable (Figure 5). Overall, however, the unequal disease duration did not explain the largely non-significant group comparisons across the analyzed domains.

**Figure 5:**
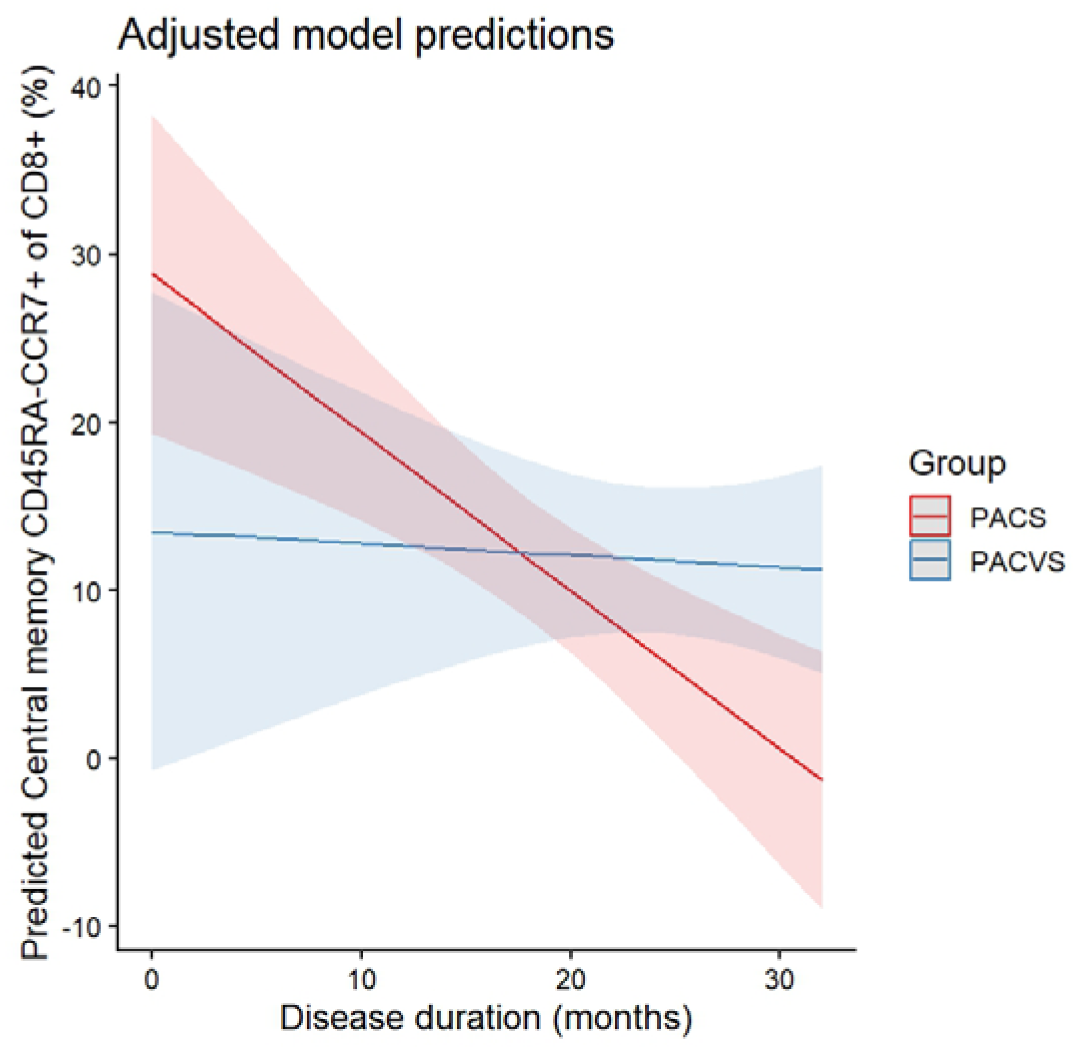
Adjusted model predictions for central memory CD45RA-CCR7+ CD8+ T-cells across disease duration. Predicted proportions of central memory CD45RA-CCR7+ CD8+ T-cells (% of total CD8+ T-cells) are shown as a function of disease duration (months) for PACS and PACVS, based on a linear regression model adjusted for age and sex and including group-by-disease-duration interaction term. Solid lines represent model-based estimates, and shaded areas indicate 95% confidence intervals. The model revealed a significant interaction after false discovery rate (FDR) correction, with a declining trajectory over time in PACS and a comparatively stable pattern in PACVS.

## 4. Discussion

In this cross-sectional study, we directly compared clinical and immunological characteristics of patients with PACS and PACVS. The two patient groups showed substantial overlap across clinical manifestations, conventional T-cell phenotypes, GPCR-directed autoantibodies, and antigen-specific cellular immune responses. Differences between PACS and PACVS were comparatively limited, whereas both patient groups showed more pronounced alterations relative to convalescent controls. These findings suggest that PACS and PACVS may share downstream immune-mediated features, although the mechanisms underlying these alterations remain to be established.

Clinically, both cohorts were characterized by prominent fatigue, post-exertional malaise (PEM), cognitive impairment, and other neurocognitive and autonomic symptoms. Several symptoms, including dizziness/lightheadedness, exhaustion, load-dependent fatigue, stress intolerance, memory or word-finding difficulties, mental exhaustibility, noise sensitivity, and sore throat, were more prevalent or severe in PACS. Nevertheless, overall functional impairment assessed by the Bell Score was comparable between PACS and PACVS. This pattern is consistent with previous studies describing substantial symptomatic overlap between PACS, PACVS, and ME/CFS (9, 27–35). Together, these findings indicate a broadly shared clinical phenotype despite differences in the severity of selected symptoms.

One of the most notable shared immunological findings was GPCR-directed autoreactivity. Autoantibodies against β1- and β2-adrenergic receptors were among the most prevalent in both PACS and PACVS, with β2-adrenergic receptor autoantibody concentrations being significantly increased compared with convalescent controls. GPCR-directed autoantibodies have previously been reported in PACS and have been linked to autonomic and cardiovascular dysfunction (12, 20–22, 36–38). However, their functional relevance remains incompletely understood, particularly because GPCR autoantibodies may also occur outside overt disease. Our observation of highly similar autoantibody profiles in PACS and PACVS nevertheless supports the presence of a shared pattern of autoreactivity.

Several mechanisms have been proposed to explain the development of autoreactivity following SARS-CoV-2 infection or vaccination, including molecular mimicry, anti-idiotypic immune responses, and persistent inflammatory immune activation (37, 39–53). These mechanisms remain hypothetical in the context of the present study and cannot be distinguished by our cross-sectional design.

Importantly, GPCR-directed autoantibodies were accompanied by cellular responses against related receptor antigens. β1/β2- and M3/M4-reactive CD4+ T cells were significantly increased in both PACS and PACVS compared with convalescent controls. Phenotypic alterations within these populations involved markers associated with activation, differentiation, and inhibitory regulation, including CTLA-4, CXCR5, LAG-3, and PD-1, together with altered naïve, central-memory, effector-memory, and TEMRA distributions. Autoreactive T cells can also be detected in healthy individuals, where they are usually controlled by peripheral tolerance mechanisms (54–57). Thus, the presence of receptor-reactive T cells alone does not establish pathogenic autoimmunity. Nevertheless, the convergence of humoral and cellular reactivity toward related receptor targets is compatible with receptor-directed immune dysregulation and warrants further mechanistic investigation.

In contrast to receptor-reactive T-cell responses, Spike-specific CD4+ T-cell frequencies were lower in PACS and PACVS than in convalescent controls. This differs from several earlier studies reporting persistent or increased SARS-CoV-2-specific T-cell responses following infection (58–62). Differences in the interval between antigen exposure and sampling may contribute to these divergent findings, as antigen-specific responses evolve over time and may be affected by repeated antigen exposure, functional exhaustion, or redistribution between blood and tissues (63–67). These explanations remain speculative because exposure history and tissue-resident immune responses were not comprehensively assessed in the present study.

Circulating Spike protein was detected in both patient cohorts, with a numerically higher prevalence in PACVS than in PACS, although the difference between the patient groups was not statistically significant. Persistent circulating or tissue-associated SARS-CoV-2 antigens have previously been reported in subsets of patients following infection (12, 18, 19, 66, 68). Data on persistent Spike following vaccination are considerably more limited (23, 69). The biological relevance of detectable Spike remains uncertain, and the present study cannot determine its origin or whether it contributes causally to persistent symptoms. Chronic antigen exposure could theoretically contribute to prolonged immune stimulation, but this requires confirmation in longitudinal and mechanistic studies (63, 70, 71).

Conventional immunophenotyping further demonstrated that PACS and PACVS were immunologically more similar to each other than either was to convalescent controls. Only three of 57 parameters differed significantly between PACS and PACVS in the unadjusted analyses, whereas both patient groups showed multiple alterations relative to controls, particularly within CD4+ and memory T-cell compartments. These findings partially align with previous reports describing changes in naïve, effector-memory, and central-memory T-cell populations in PACS (72–75).

EBV reactivation did not appear to represent a major distinguishing feature in our cohorts. Detectable EBV DNA was uncommon and did not differ significantly between PACS and PACVS. Previous studies have reported associations between EBV reactivation and PACS, although prevalence estimates vary according to the diagnostic method and biological material analyzed (76–80). Our blood-based qPCR approach may underestimate previous or compartmentalized EBV reactivation, as serological markers or salivary EBV DNA may identify additional cases. Accordingly, our findings do not exclude a role for EBV in individual patients but provide no evidence that systemic EBV reactivation represents a common mechanism distinguishing PACS from PACVS.

Taken together, our findings demonstrate substantial clinical and immunological overlap between PACS and PACVS. Particularly notable were the shared certain GPCR autoantibody profiles, increased β1/β2- and M3/M4-reactive CD4+ T-cell responses, and similar alterations in conventional T-cell phenotypes relative to convalescent controls. The convergence of humoral and cellular autoreactivity toward receptors involved in autonomic regulation is compatible with an immune-mediated component in both conditions. However, the present data cannot establish whether these alterations are pathogenic drivers, consequences of persistent immune activation, or secondary disease markers.

### 4.1 Limitations

Several limitations should be considered. The first and most important limitation concerns the etiological classification of patients with PACVS. Currently, no established biomarker or immunological signature can reliably distinguish PACVS from PACS at the individual-patient level. Therefore, an unrecognized or asymptomatic SARS-CoV-2 infection preceding symptom onset cannot be excluded with certainty in patients classified as PACVS, and some manifestations attributed to vaccination could theoretically represent infection-associated PACS. This is particularly relevant given the substantial clinical and immunological overlap observed between the two syndromes and warrants cautious interpretation of our findings. Nevertheless, patients classified as PACVS reported symptom onset in close temporal association with COVID-19 vaccination and, according to the available clinical history, no symptomatic acute COVID-19 episode or known positive SARS-CoV-2 test temporally associated with syndrome onset. These clinical characteristics support vaccination as the presumed trigger; however, they cannot definitively establish causality or exclude an unrecognized preceding infection.

Further, the cross-sectional observational design precludes conclusions regarding causality or the temporal development of the observed immune alterations. The relatively small cohort size limits statistical power and increases uncertainty around subgroup comparisons. Furthermore, due to the random patient recruitment, several baseline characteristics including age, sex and duration since the last antigenic contact differed between the groups and represent another potential limitation. However, multivariate analyses did not identify any effect of these factors on our main results.

Symptom assessments were based predominantly on patient-reported questionnaires and may therefore be affected by reporting bias.

In conclusion, PACS and PACVS showed substantial clinical and immunological similarities despite different preceding antigen exposures. Compared with convalescent controls, both patient groups exhibited increased concentration of β-adrenergic receptor directed autoantibodies, increased β1/β2- and M3/M4-reactive CD4+ T-cell responses, altered conventional T-cell phenotypes, and detectable circulating Spike protein in subsets of patients. Differences between PACS and PACVS were comparatively limited. These findings identify shared immune alterations and support further investigation of immune-mediated and potentially autoreactive mechanisms in both conditions. Larger, prospectively characterized and longitudinal cohorts are required to validate these observations, determine their relationship to clinical manifestations, and establish whether the identified immune alterations contribute causally to disease persistence.

## Data availability statement

The data can be provided upon special request to the authors.

## Author contributions

Data analysis: JD, KSR, MA, EK, JK, US, SS Recruitment: JD, LW, FS, THW, NB, Manuscript: JD, KSR, NB

## Acknowledgements

We sincerely thank all patients and participants for their valuable contribution to this study

## Abbreviations

AT-II: Angiotensin II receptor type 1
BSA: Bovine serum albumin
CCC: Canadian consensus criteria questionnaire
CCG: Convalescent control group
CM: Central-memory T cells
DMSO: Dimethyl sulfoxide
EBV: Epstein-Barr virus
EM: Effector-memory T cells
ET-A: Endothelin A receptor
FCS: Fetal calf serum
FDR: False Discovery Rate
GPCR: G-protein-coupled receptor
M3: muscarinic acetylcholine receptor
M3 M4: muscarinic acetylcholine receptor M4
ME/CFS: Myalgic encephalomyelitis/chronic fatigue syndrome
PACS: Post acute COVID-19 syndrome
PACVS: Post acute COVID-19 vaccination syndrome
PBMCs: Peripheral blood mononuclear cells
PBS: Phosphate-buffered saline
PEM: Post-exertional malaise
SEB: Staphylococcal enterotoxin B
SSSQ: Subjective symptom severity questionnaire
β1: β1-adrenergic receptor
β2: β2-adrenergic receptor

