## Supplemental Figures for "Shared Clinical and Immunological Features of Post-Acute COVID-19 and Post-Acute COVID-19 Vaccination Syndromes"

### Supplementary Information

Supplemental Table S1: Cohort characteristics and measured tests. *n* = number, SD = standard deviation

| | PACS | PACVS | Ctrl |
|---|---|---|---|
| Patients, *n* | 28 | 29 | 16 |
| Age (mean ± SD) | 48.21 ± 13.43 | 39.97 ± 11.08 | 37.56 ± 12.52 |
| Female sex, *n* (%) | 25 (89.3) | 16 (55.2) | 10 (62.5) |
| ME/CFS, *n* (%) | 21 (75) | 18 (62) | - |
| **Tests measured** | | | |
| Immunophenotyping, *n* | 28 | 29 | 16 |
| Autoantibodies, *n* | 27 | 29 | 16 |
| Spike, *n* | 25 | 28 | 16 |
| EBV, *n* | 25 | 26 | 16 |
| Ag. spec. TC, *n* | 19 (18 M3/M4) | 19 (20 Spike) | 16 |

Supplemental Table S2: Time from antigen contact until measurement of the different tests in months. The numbers displayed show the median ± standard deviation.

| | **PACS** | **PACVS** |
|---|---|---|
| Immunophenotyping (mean ± SD) | 18.22 ± 7.23 | 22.97 ± 6.18 |
| Autoantibodies (mean ± SD) | 19.08 ± 6.72 | 23.79 ± 6.10 |
| Spike (mean ± SD) | 21.17 ± 7.74 | 25.46 ± 6.71 |
| EBV (mean ± SD) | 18.92 ± 6.72 | 23.62 ± 6.16 |
| Ag. spec. TC (mean ± SD) | 23.05 ± 11.06 | 29.79 ± 6.41 |

Supplemental Table S3: Symptom frequencies reported in the subjective Canadian Consensus Criteria questionnaire. *ns* = not significant.

| | PACS (% of 27) | PACVS (% of 26) | *p*-values |
|---|---|---|---|
| **New physical/mental exhaustion** | 77.78 | 76.92 | *ns* |
| **Exhaustion with delayed recovery** | 85.19 | 73.08 | *ns* |
| **Deterioration due to exertion/stress** | 100.00 | 96.15 | *ns* |
| **Difficulty falling asleep** | 59.26 | 53.85 | *ns* |
| **Sleep disturbances** | 70.37 | 57.69 | *ns* |
| **Change in day/night rhythm** | 33.33 | 26.92 | *ns* |
| **Restless sleep** | 85.19 | 73.08 | *ns* |
| **Joint pain** | 55.56 | 53.85 | *ns* |
| **Muscle pain** | 85.19 | 61.54 | *ns* |
| **Headache** | 81.48 | 65.38 | *ns* |
| **Concentration impairment** | 96.30 | 88.46 | *ns* |
| **Problems processing information** | 81.48 | 76.92 | *ns* |
| **Perceptual & sensory disorders** | 40.74 | 46.15 | *ns* |
| **Disorientation** | 22.22 | 26.92 | *ns* |
| **Problems with coordination of movement** | 37.04 | 26.92 | *ns* |
| **Symptoms of overload** | 96.30 | 76.92 | *ns* |
| **Dizziness during rapid changes in position** | 77.78 | 65.38 | *ns* |
| **POTS** | 29.63 | 50.00 | *ns* |
| **Dizziness/lightheadedness** | 70.37 | 34.62 | 0.0135 |
| **Extreme paleness** | 18.52 | 19.23 | *ns* |
| **Intestinal dysfunction** | 59.26 | 65.38 | *ns* |
| **Bladder dysfunction** | 25.93 | 23.08 | *ns* |
| **Vasomotor instability** | 29.63 | 23.08 | *ns* |
| **Dyspnea** | 51.85 | 38.46 | *ns* |
| **Disturbed body temperature regulation** | 40.74 | 26.92 | *ns* |
| **Heat intolerance** | 74.07 | 57.69 | *ns* |
| **Cold intolerance** | 33.33 | 15.38 | *ns* |
| **Loss of appetite** | 37.04 | 57.69 | *ns* |
| **Loss of weight** | 22.22 | 7.69 | *ns* |
| **Low blood sugar** | 0 | 11.54 | *ns* |
| **Poor stress management** | 88.89 | 80.77 | *ns* |
| **Stress increases exhaustion** | 33.33 | 19.23 | *ns* |
| **Painful lymph nodes** | 25.93 | 26.92 | *ns* |
| **Recurrent sore throat** | 59.26 | 34.62 | *ns* |
| **New allergies/changes** | 25.93 | 30.77 | *ns* |
| **Flu-like symptoms** | 81.48 | 61.54 | *ns* |
| **Hypersensitivity to medications** | 37.04 | 30.77 | |

Supplemental Table S4: Subjective symptom severity reported in the questionnaire of the study group. Values are reported as mean values with interquartile range, unless otherwise noted. For the following points the number of events were analyzed instead of the severity: Infections UAW in last 12 M, infections LAW in last 12 M, Infections Herpes in last 12 M, Other infections in last 12 M and Antibiotics in last 12 M. *ns* = not significant.

| | PACS | PACVS | *p*-values |
|---|---|---|---|
| **Exhaustion** | 8.00 [4-10] | 8.00 [2-10] | 0.0327 |
| **Load-dependent increase in fatigue the next day** | 9.00 [4-10] | 7.00 [2-10] | 0.0023 |
| **Need for rest breaks** | 8.00 [1-10] | 8.00 [5-10] | *ns* |
| **Limitations in performance in everyday life** | 8.00 [3-10] | 8.00 [4-10] | *ns* |
| **Intolerance of stress** | 8.50 [1-10] | 8.00 [1-10] | 0.0153 |
| **Muscle pain** | 5.50 [0-10] | 6.00 [1-10] | *ns* |
| **Headache** | 6.00 [1-10] | 5.00 [1-10] | *ns* |
| **Joint pain** | 3.00 [1-10] | 5.00 [1-10] | *ns* |
| **Difficulties with memory/word-finding** | 7.00 [1-10] | 5.00 [1-9] | 0.0493 |
| **Difficulty concentrating** | 8.00 [2-10] | 6.00 [1-10] | *ns* |
| **Mental exhaustibility** | 8.00 [3-10] | 6.00 [1-9] | 0.0158 |
| **Visual impairment** | 2.00 [1-10] | 5.00 [1-10] | *ns* |
| **Mood swings** | 4.50 [1-10] | 4.50 [1-8] | *ns* |
| **Reading comprehension** | 6.50 [1-10] | 6.00 [1-10] | *ns* |
| **Palpitations** | 3.00 [1-10] | 4.00 [1-10] | *ns* |
| **Dizziness when standing up** | 5.00 [1-10] | 5.00 [1-10] | *ns* |
| **Dizziness when walking** | 4.00 [1-10] | 3.50 [1-10] | *ns* |
| **Sleep disturbances** | 8.00 [1-10] | 6.00 [1-10] | *ns* |
| **Temperature sensitivity** | 6.00 [1-10] | 6.50 [1-10] | *ns* |
| **Light sensitivity** | 5.50 [1-10] | 5.00 [1-10] | *ns* |
| **Noise sensitivity** | 7.50 [1-10] | 5.00 [1-9] | 0.0085 |
| **Dyspnea** | 3.50 [1-9] | 2.50 [1-10] | *ns* |
| **Irritable bowel** | 5.00 [1-10] | 6.00 [1-10] | *ns* |
| **Fever** | 1.00 [1-6] | 1.00 [1-8] | *ns* |
| **Painful lymph nodes** | 1.00 [1-8] | 1.00 [1-10] | *ns* |
| **Sore throat** | 3.50 [1-10] | 1.50 [1-10] | 0.0319 |
| **Flu-like symptoms** | 5.50 [1-10] | 5.00 [1-10] | *ns* |
| **Infections UAW in last 12 M** | 1.38 [0-8] mean | 2.96 [0-20] mean | *ns* |
| **Infections LAW in last 12 M** | 0.42 [0-3] mean | 1.18 [0-20] mean | *ns* |
| **Infections Herpes in last 12 M** | 1.62 [0-12] mean | 0.89 [0-9] mean | *ns* |
| **Other infections in last 12 M** | 1.12 [0-17] mean | 0.61 [0-5] mean | *ns* |
| **Antibiotics in last 12 M** | 0.65 [0-6] mean | 0.61 [0-5] mean | *ns* |
| **Self-assessment of disease severity** | 8.00 [3-10] | 8.00 [5-10] | *ns* |

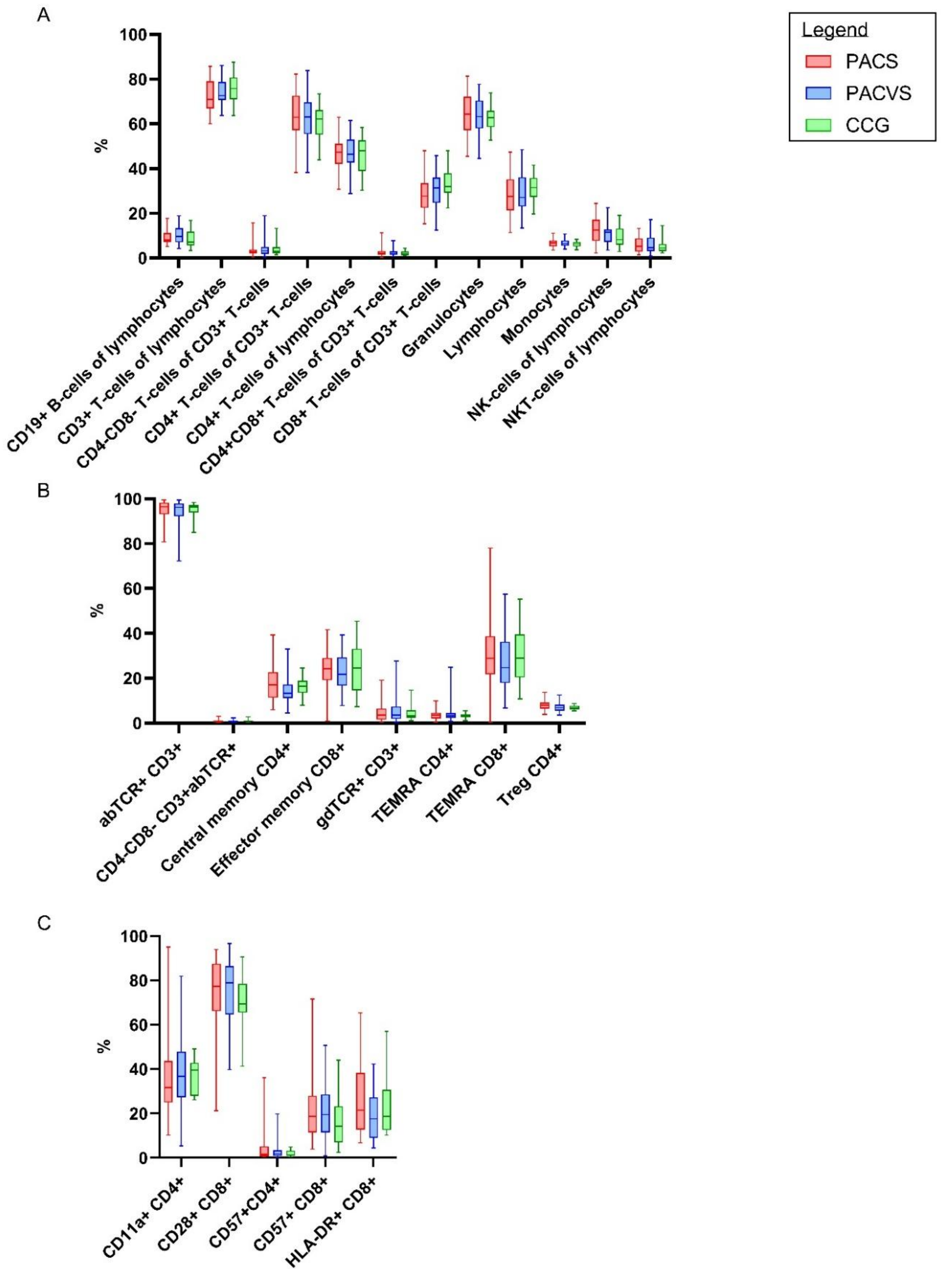


Supplemental Figure S1: **(A)**: Immunophenotyping analysis, comparison of the subgroup basic immune status. **(B)**: Immunophenotyping analysis, comparison of the subgroup α/β TmemTreg. **(C)**: Immunophenotyping analysis, comparison of the subgroup TC act. Statistical analyses performed by Mann-Whitney U-test with the following p-values: * = $p<0.05$, ** = $p<0.01$, *** = $p<0.001$, **** = $p<0.0001$.

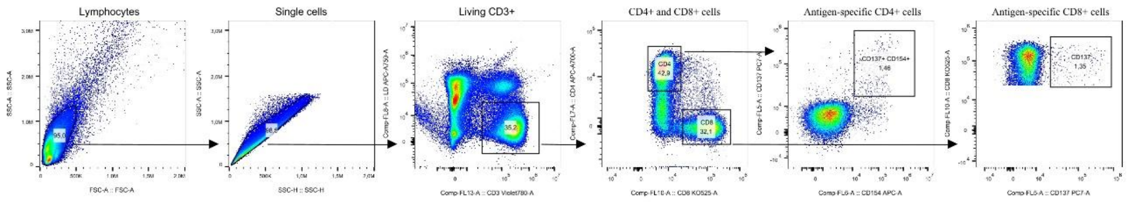


Supplemental Figure S2: Gating strategy for the identification of antigen-specific T-cells. Identification of lymphocytes with SSC/FSC profile. Inclusion of single cells, followed by identification of the living CD3$^+$. CD3$^+$ cells were split up into CD4$^+$ and CD8$^+$, from which antigen-specific T-cells were identified.

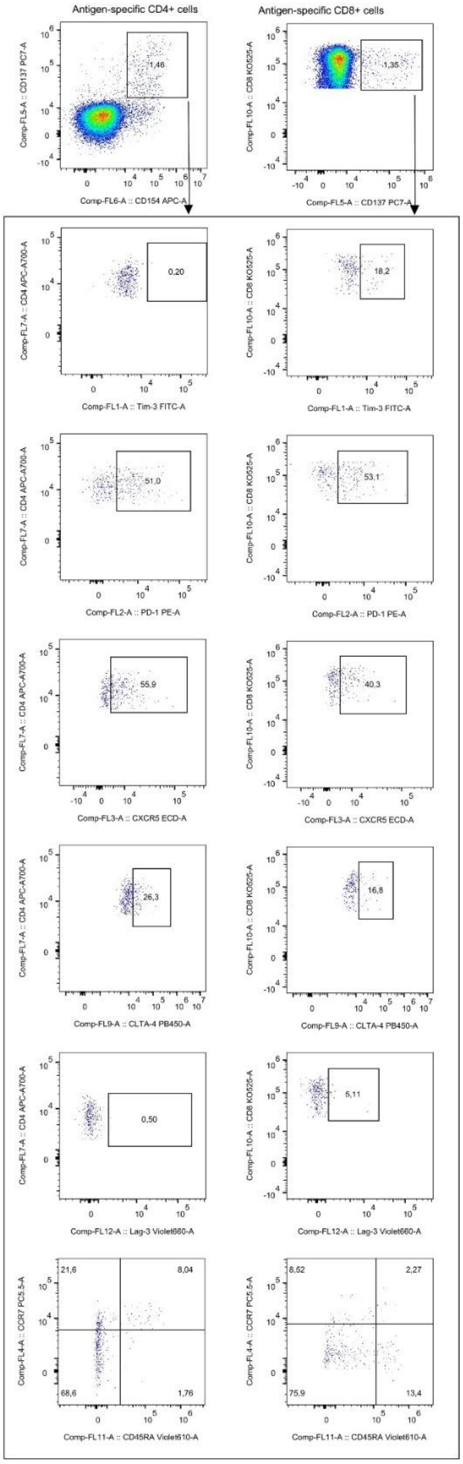


Supplemental Figure S3: Gating strategy to identify subpopulations with Spike protein and GPCR-receptor specificity. After stimulation of PBMC with different antigens (ß1/ß2 receptor lysates, M3/M4 receptor lysates, Spike protein OPPs. Identification of antigen specific T-helper cells ($CD4^+$ $CD137^+$ $CD154^+$) and antigen specific cytotoxic cells ($CD8^+$ $CD137^+$). Cells were stained for the exhaustion markers Tim-3, PD-1, CTLA-4, Lag-3 and the chemokine receptor CXCR5. Naïve antigen specific T-cells ($CD45RA^+$ $CCR7^+$), central memory T-cells ($CD45RA^-$ $CCR7^+$), effector memory T-cells ($CD45RA^-$ $CCR7^-$) and TEMRA cells ($CD45RA^+$ $CCR7^-$) were identified.

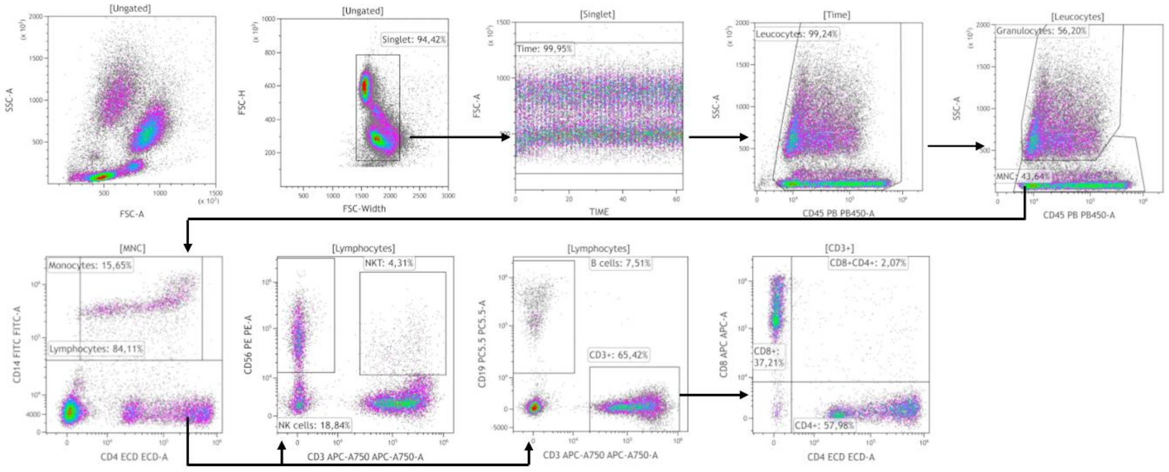


Supplemental Figure S4: Gating strategy basic immune status. Mononuclear cells and granulocytes were identified by side scatter profile and CD45 expression. Mononuclear cells were separated in monocytes [$CD14^{+}$) and lymphocytes [$CD14^{-}$). Lymphocytes were separated into NK [$CD3^{-}$ $CD56^{+}$), NKT [$CD3^{+}$ $CD56^{+}$). T-cells and B-cells were identified by CD3 and CD19 and T-cells were then distinguished into $CD4^{+}$ helper T-cells and $CD8^{+}$ cytotoxic T-cells.

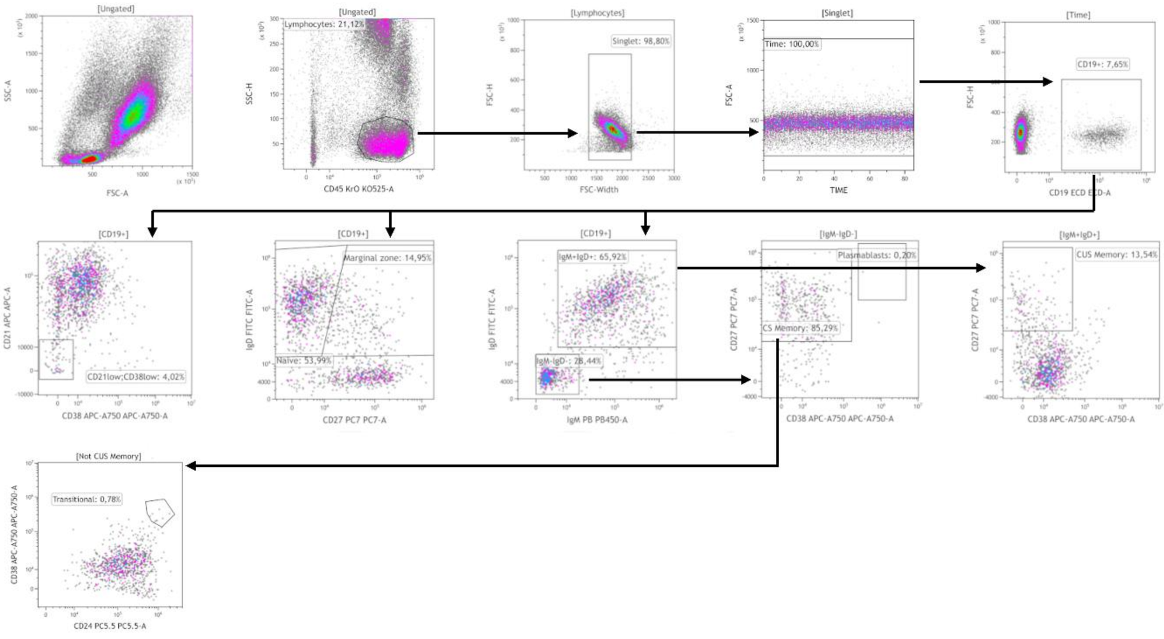


Supplemental Figure S5: Gating strategy B-cells: Single lymphocytes were identified with their side scatter profile and CD45. B-cells were identified [$CD19^+$). B-cell subpopulations were identified: naïve B-cells [$IgD^+$ $CD27^-$, non-switched memory [$CD38^-$ $CD27^+$ $IgM^-$ $IgD^-$), Plasmablasts [$CD38^+$ $CD27^+$), switched memory [$CD27^+$ $CD38^-$ $IgM^+$ $IgD^+$), traditional B memory cells [$CD38^+$ $CD24^+$) and $CD21l^{ow}CD38^{low}$ B-cells.

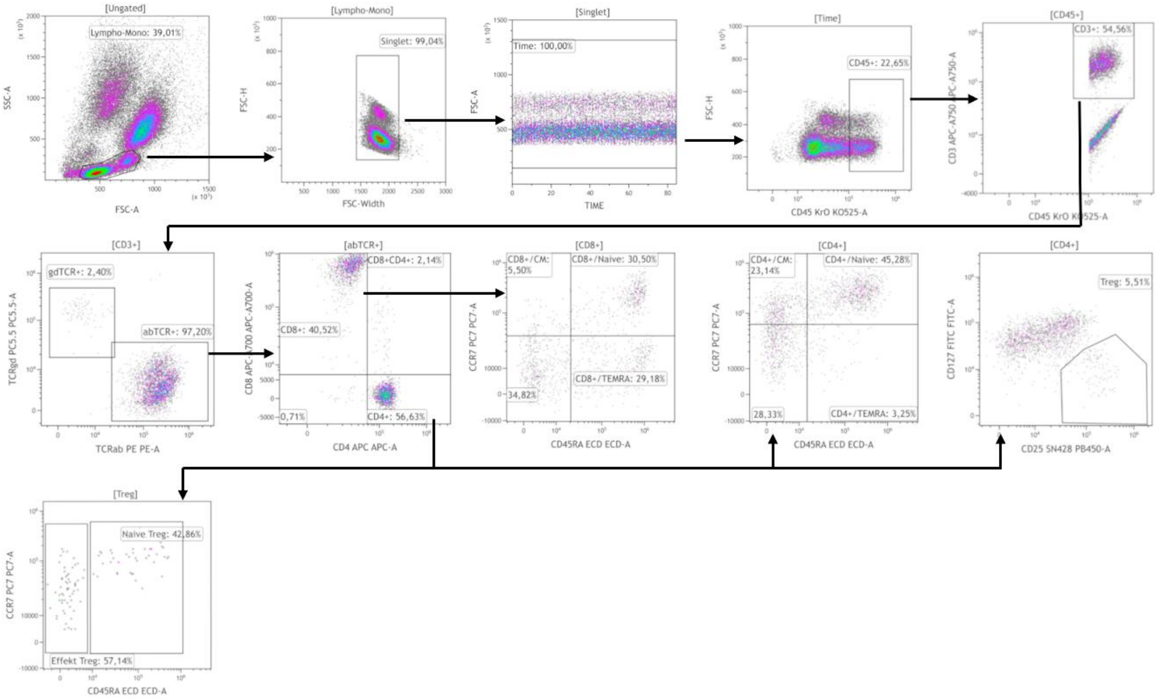


Supplemental Figure S6: Gating strategy T-memory and Tregs: T-lymphocytes were identified with CD45 and CD3, then subdivided by their expression of α/β T-cell receptor and γ/δ T-cell receptor. αβ$TCR^+$ cells were divided into memory $CD4^+$, memory $CD8^+$ cells, Tregs. Memory $CD4^+$ were identified: central memory [$CD45RA^-$ $CCR7^+$), naïve [$CD45RA^+$ $CCR7^+$), effector memory [$CD45RA^-$ $CCR7^-$) and TEMRA [$CD45RA^+$ $CCR7^-$). Tregs were divided into naïve Tregs [$CD45RA^+$) and effector Tregs [$CD45RA^-$).

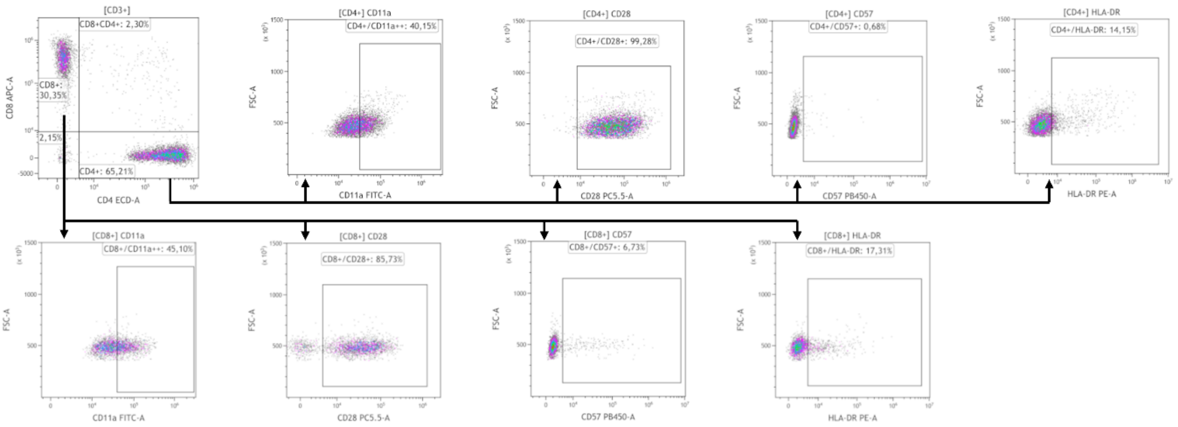


Supplemental Figure S7: Gating strategy T-cell activation: $CD3^{+}$ cells were identified and divided into $CD4^{+}$ and $CD8^{+}$ T-cells. Activated T-cells were identified through expression of different activation markers [CD11a, CD28, CD57, HLA-DR).
